# Implementation of digital chest radiography for childhood tuberculosis diagnosis at district hospital level in six high tuberculosis burden and resources limited countries

**DOI:** 10.1101/2024.08.23.24312489

**Authors:** Bernard Fortune Melingui, Basant Joshi, Jean-Voisin Taguebue, Douglas Mbang Massom, Etienne Leroy Terquem, Pierre-Yves Norval, Angelica Salomao, Dim Bunnet, Tek Chhen Eap, Laurence Borand, Celso Khosa, Raoul Moh, Juliet Mwanga-Amumpere, Mao Tan Eang, Ivan Manhiça, Ayeshatu Mustapha, Eric Balestre, Samuel Beneteau, Eric Wobudeya, Olivier Marcy, Joanna Orne-Gliemann, Maryline Bonnet, the TB-Speed Decentralization study group

**Author notes:** Corresponding author Bernard Fortune MELINGUI Translational Research on HIV and Endemic and Emerging Infectious Diseases (TransVIHMI), University of Montpellier, National Institute for Health and Medical Research (INSERM), French National Research Institute for Sustainable Development (IRD), Montpellier, France 911 avenue Agropolis, 34394 Montpellier cedex 5, France.

## Abstract

**Objectives:** Chest X-ray (CXR) plays an important role in childhood tuberculosis (TB) diagnosis but access to quality CXR remains a major challenge in resource-limited settings. Digital CXR (d-CXR) can solve some image quality issues and facilitate their transfer for quality control. We describe the implementation of introducing d-CXR in twelve district hospitals (DH) in 2021-22 across Cambodia, Cameroon, Ivory Coast, Mozambique, Sierra Leone and Uganda as part of the TB-Speed decentralization study on childhood tuberculosis diagnosis.

**Methods:** For digitization of CXR Digital Radiography (DR) plates was set-up on existing analogue radiography machines. D-CXR were transferred to an international server at Bordeaux University and downloaded by sites’ clinicians for interpretation. We assessed the pre-intervention (baseline situation and d-CXR set-up) and per-intervention (uptake, challenges and health care workers’ (HCW) perceptions) of d-CXR implementation. We used a convergent mixed method approach utilizing process data, individual interviews with 113 HCWs involved in performing or interpreting d-CXRs and site support supervision reports.

**Results:** Of 3104 children with presumptive TB, 1642 (52.9%) had at least one d-CXR including 1505, 136 and 1 children with one, two and three d-CXR respectively, resulting in a total of 1780 d-CXR. Of them, 1773 (99.6%) were of good quality and 1772/1773 (99.9%) were interpreted by sites’ clinicians. 164 children had no d-CXR performed despite attending the radiography department: 126, 37 and 1 with one, two and three attempts, respectively. D-CXRs were not performed in 21.6% (44/203) due to connectivity problem between the DR plate captor and the computer. HCWs reported good perceptions of d-CXR and of the DR plates provided. The main challenge was the upload to and download from the server of d-CXRs, due to limited internet access.

**Conclusion:** D-CXR using DR plates was feasible at district hospital level and provided good quality images but required overcoming operational challenges.

## INTRODUCTION

Tuberculosis (TB) is one of the leading infectious causes of death worldwide in children with 183 000 deaths in 2022 (1). Modelling suggest that more than 95% of these children were not treated hence likely not diagnosed at the time of death (2).

Microbiological confirmation of childhood TB is difficult due to challenges in obtaining respiratory samples in young children and the paucibacillary nature of TB in children resulting in low sensitivity of bacteriological tests from respiratory samples (3,4). Consequently, the majority of children are clinically diagnosed for TB, mainly based on clinical evaluation and chest X-ray (CXR) findings (5). CXR plays a critical place in the recent treatment decision algorithm proposed by WHO for tuberculosis in children (6,7).

In high TB burden and limited resources countries, the use of CXR faces several difficulties: i) radiology services are often only available in district or regional hospitals (8), hence requiring referral for CXR of children with presumptive TB seen at Primary Health Centres (PHC), inducing additional transport costs and affecting access to CXR; ii) clinicians lack training in interpreting CXR for paediatric TB diagnosis, especially at peripheral healthcare facilities affecting the reliability of CXR results; and iii) films and reagents for analogue X-ray machines often suffer from shortage and expiries, leading to poor CXR quality (9).

Digital CXR (d-CXR) could contribute to improving the quality of CXR interpretation in resource-limited settings by overcoming the issue of shortage and poor quality films and reagents (10). D-CXR could also allow remote interpretation for advice or external reading for quality control purpose or advice. In addition, digital technologies can enable future use of computer aid detection (CAD) for TB, already validated for adults, and under evaluation in children (11). The implementation of d-CXR for childhood TB diagnosis has been poorly documented in resource-limited settings. Documenting and learning from the challenges and solutions found in operational settings is essential to inform implementers and decision makers on the use of d-CXR at lower levels of care. Using the experience of the TB-Speed decentralization study, we described the implementation of d-CXR in 12 DHs of six high TB incidence and resource-limited settings.

## METHODS

### Study setting

The TB-Speed decentralization study was an operational research to assess the effect of implementing a comprehensive diagnostic approach on paediatric TB case detection at DH and PHC levels. The approach included symptomatic TB screening of all sick children (<15 years) arriving at the health facility to identify children with presumptive TB, clinical evaluation for all children with presumptive TB, microbiological testing using Xpert MTB/RIF Ultra on a nasopharyngeal aspirate (NPA) and a stool or sputum sample, and d-CXR. The study was implemented in two rural or semi-rural health districts in Cambodia, Cameroon, Ivory Coast, Mozambique, Sierra Leone and Uganda (12)). In one district, the DH implemented the diagnosis approach, while PHCs only referred children identified with presumptive tuberculosis to DH after screening. In the other district both the DH and PHCs implemented the diagnosis approach, except for CXR that was available at DH only. The study comprised an observation period to document baseline situation followed by a lead-in phase to set-up and build capacity (pre-intervention), and a 12 months of intervention period (per-intervention).

### Digital radiography set up and supervision

For DH without existing digital CXR system, analogue radiographies were equipped with digital radiography (DR) plates (Agfa DR14eC, AGFA N.V, France), which produces a digital radiographic image instantly on a computer. For DH without functional radiography, a portable radiography was provided. Sites received support to strengthen internet connexion if needed and to reinforce protective measures when necessary. Sites’ radiographers were trained on how to convert Digital Imaging and Communication in Medicine (DICOM) images’ format into Joint Photographic Experts Group (JPEG) format and to upload them on the international FTPS (File Transfer Protocol Secure) server at the University of Bordeaux (France). Sites’ clinicians and radiographers were trained on how to download d-CXR images on tablets for interpretation and on assessing the quality of CXR using standardised criteria as part of a 1.5-day training course (13). CXR were performed using antero-posterior and lateral CXR for children younger than 5 years and postero-anterior CXR for children older than 5 years. If a CXR was assessed as uninterpretable due to poor quality, another CXR was performed immediately. The interpretation was recorded by readers in an electronic Case Report Form (CRF) data on REDCap clinical database on tablets. Support supervision activities were implemented to support field implementation of the diagnostic approach including CXR with visits conducted by representative of national TB programs (NTP), paediatricians and study country coordinators, guided by a standardised support supervision tool documenting both quantitative and qualitative data related to performance, challenges observed and/or discussed and immediate actions taken or recommendations made for further action.

### Implementation study design and population

Within the TB-Speed Decentralization study, we conducted a broad implementation research programme, using a convergent mixed methods design. The pre-intervention component of the implementation of d-CXR relied on baseline X-ray capacity and setup data, and the per-intervention component on process data, individual interviews data and support supervision data. Qualitative and quantitative baseline capacity and setup data were retrieved from baseline assessment forms and from country implementation guide, respectively. Quantitative process data were obtained from the clinical database for children with presumptive TB enrolled in the TB-Speed Decentralization study. These children were recruited between August 2019 and September 2021. Qualitative interviews were conducted among HCWs who were either directly involved in performing the d-CXR (radiographers), and/or who read and interpreted images (medical doctors, nurses) and/or who took part in other components of the diagnosis approach, such as screening, and observed the d-CXR process (midwifes and paramedics). Support supervision quantitative and qualitative data was retrieved from site supervision reports.

### Outcomes

#### Pre-intervention

Pre intervention d-CXR implementation outcomes consisted first in baseline radiography capacity in each DH: availability of an X-Ray unit with X-ray protection, functional analogue X-ray machine, availability of digital X-Ray, stabilising and back-up systems for electricity and number of radiographers (14). Other outcome related to the level of capacity strengthening for the d-CXR setup in terms of renovation of the X-ray unit, number and type of equipment provided, additional logistic support with electricity and number of radiographers trained for equipment use (Table 1).

**Table 1:** Summary of the analytical approach.

| Objectives | Endpoints | Endpoint measures | Population | Primary source of information |
| --- | --- | --- | --- | --- |
| To describe the pre-intervention implementation of d-CXR | Baseline d-CXR capacity | - Availability of X-ray ward<br>- Availability of analogic functional X-ray machine<br>- Availability of digital X-ray<br>- Availability of electricity<br>- Number of radiographers available |  | Pre-intervention site assessment report |
|  | Set-up of d-CXR | - Logistic support<br>- Provision of X-ray machine<br>- Provision of DR plate and accessories<br>- Number of radiographers trained<br>- Provision in internet |  | Country Implementation guide |
| To assess the per-intervention implementation of d-CXR | CXR performance uptake and interpretation by clinician at facility | - Number of children with presumptive TB<br>- Number of children with CXR performed<br>- Number of CXR performed per child<br>- Number of children with CXR attempts | Children with presumptive TB enrolled in the TB-Speed decentralisation study | REDCap clinical Database |
|  |  | - Number of CXR attempts<br>- Number and proportion of CXR performed<br>- Number and proportion of good quality CXR<br>- Number and proportion of interpreted CXR | CXR attempts at district hospital level | REDCap clinical Database |
|  | Reasons for CXR not performed | - Technical problem with the X-ray machine<br>- Problems of connectivity between with the DR plate<br>- Electrical problems<br>- Other technical problems<br>- Absence of radiographer<br>- Administrative issues | CXR attempts without any CXR performed | REDCap clinical Database |
|  | d-CXR performance indicators | - Median score of d-CXRs performed as indicated<br>- Median score of d-CXR loaded on/from the server as indicated<br>- Median score of d-CXRs interpreted as indicated | Supervisors | REDCap database of site supervision reports |
|  | d-CXR challenges and recommendations identified by supervisors | - Loading of d-CXR<br>- Technical problem with DR plates<br>- Issues with internet connection<br>- Delays in d-CXR interpretation<br>- Quality issues<br>- Dysfunction of the X-ray machine | Supervisors | REDCap database of site supervision reports |
|  | Perception of d-CXR by health care workers | Experience and views of health care workers regarding d-CXR | HCWs involved in diagnosis of tuberculosis in children | Semi-structured interviews |

PHC: Primary health Centre; DH: District Hospital; CXR: Chest X- ray; TB: Tuberculosis; HCW: Healthcare worker

#### Per-intervention

Per intervention d-CXR implementation outcomes included process outcomes, supervision outcomes and individual experiences and perceptions. Process outcomes included the uptake of d-CXR assessed by the cascade of care from the CXR prescription to its interpretation: 1) number of children with i) presumptive TB, ii) at least one d-CXR performed (one, two and three d-CXR performed), iii) any CXR attempts (visit to X-ray department) but without d-CXR performed (one, two and three attempts); 2) number of d-CXR i) performed, ii) of good quality defined as readable, and iii) interpreted; 3) reasons for not performing a d-CXR presented per category as recorded by sites’ clinicians including: technical problems with X-ray machine, connectivity problem between the DR plate detector and the computer, electrical problems, absence of radiographer, administrative reason and non-specified technical issues. Supervision outcomes assessed performance of CXR services by supervisors, through 3 quantitative indicators: 1) d-CXRs performed as prescribed, assessed by the availability of a d-CXR images for children with presumptive TB at the DH in the last 2 weeks; 2) Upload and download of d-CXR images, assessed by the ability of the site radiographer and site reader to upload and download d-CXR image; 3) d-CXRs interpretation, assessed by the presence of at least two d-CXR interpretation reports from the last 2 weeks. Performance outcomes were scored using a Likert scale as follows; 1-Never: The task is not done completely; 2-Rarely: Performance of task is below average; 3-Sometimes: Performance of task is average; 4-Often: Performance of task is above average; 5-Always. Supervision outcomes included also action taken and recommendations following supervision visits. Individual HCWs’ experiences and perception of d-CXR included operational challenges and contextual conditions that made d-CXR possible or not (15).

### Data collection and analysis

Pre-intervention implementation data was collected between January and June 2018 from paper-based standardised baseline assessment forms, filled by country investigators and entered in an Excel database. Setup data was collected between May and December 2019 from the International Project Manager’s activity monitoring file including country implementation guides.

For the per-intervention implementation data, quantitative process data were computed from real-time patient level individual data, collected during study intervention for enrolled children by site HCW assigned to the study and directly entered into an electronic CRF. Process outcomes are presented as counts, proportions, medians and interquartile ranges (IQR). Paper-based site supervision reports were entered into a REDCap dedicated database by research assistants. We calculated the median scores’ and IQR for the three CXR outcomes.

All quantitative data were analysed with the R software (version 4.2.1, R Foundation for Statistical Computing).

Regarding per-intervention implementation qualitative data, reasons for d-CXR not performed were retrieved from the REDCap clinical database. Supervision visits qualitative notes made by supervisors were extracted from site supervision reports and entered as free text into the REDCap supervision database. Challenges to perform d-CXR and mitigations strategies were grouped into global deductive thematic categories respectively.

Individual interviews were conducted by trained social science research assistants using a semi-structured guide, between June and August 2021. Interviews aimed at investigating overall experiences and perceptions of the childhood TB diagnosis approach (16), and for this paper we analyzed data related to d-CXR process. Among the 113 HCWs interviewed, median age was 36 years (IQR; 31, 41) and 64 (57%) were female. More than half of the HCWs interviewed were nurses (Table S1). Interviews were conducted face-to-face or by telephone (when transportation to the site was not possible due to COVID-19 restrictions) and were recorded if respondents permitted. Qualitative data were anonymised using numerical identification codes. A thematic analysis was performed in three steps: we first developed a codebook based on the themes that were predefined by the semi-structured interview guides. We then coded the data using the Nvivo release qualitative data management software, QSR International Private Ltd. Version 1.5, 2021. Recurring themes were identified and grouped into overarching thematic categories related to d-CXR, and later on summarized.

### Ethics statement

This study was approved by each implementing country’s National Ethics Committees, the WHO Ethical Review Board, and the National Institute of Health and Medical Research (INSERM Ethics Evaluation Committee. Individual consent was obtained from Parents/Guardians of enrolled children and the child’s assent was obtained when the child was old enough as per countries regulation and from interviewed HCWs.

## RESULTS

### Pre-intervention implementation of d-CXR

As per the baseline DH capacity assessment, 10/12 (83%) DH had an X-ray department and an analogue X-ray machine. Only 1 out of 12 DH already had a digital X-ray (Bo Government referral hospital from Sierra Leone). Electrical power instability and cuts were reported in Cambodia, Cameroon, Cote d’Ivoire and Mozambique. In all radiology departments, at least one radiographer was available to perform CXR (Table 2).

**Table 2:** Baseline digital X-ray capacity and setup of digital X-ray per district hospital and country during pre-intervention implementation.

|  |  | Baseline capacity |  |  |  |  | Set up of digital X-ray |  |  |  |  |  |
| --- | --- | --- | --- | --- | --- | --- | --- | --- | --- | --- | --- | --- |
| Countries | District referral hospital | Availability of X-ray unit | Availability of analogic functional X-ray machine | Availability of digital X-ray | Electrical power issue | Number of radiographers in X-ray ward | Electricity provided by the study | Renovation of X-ray ward | Provision of X-ray machine | Provision of DR plate and its accessories | Number of radiographers trained | Provision of internet |
| Cambodia | Batheay | Yes | Yes | No | No | 2 | No | No | No | Yes | 2 | Yes |
|  | Angroka | Yes | Yes | No | No | 2 | No | No | No | Yes | 2 | Yes |
| Cameroon | Obala | Yes | Yes | No | No | 2 | No | No | No | Yes | 2 | Yes |
|  | Bafia | Yes | Yes | No | No | 2 | No | No | No | Yes | 2 | Yes |
| Ivory coast | Sassandra | Yes | Yes | No | Yes | 3 | No | No | No | Yes | 2 | Yes |
|  | Danane | Yes | Yes | No | No | 5 | No | No | No | Yes | 5 | Yes |
| Mozambique | Manjacaze | Yes | Yes | No | No | 3 | No | No | No | Yes | 2 | Yes |
|  | Chokwe | Yes | Yes | No | No | 2 | No | No | No | Yes | 2 | Yes |
| Sierra Leone | Bo | Yes | Yes | Yes | Yes | 5 | No | No | No | No | 5 | Yes |
|  | Port Loko | Yes | Yes | No | Yes | 2 | No | No | No | Yes | 2 | Yes |
| Uganda | Rakai | No | No* | No | Yes | 1 | No | Yes | Yes | Yes | 1 | Yes |
|  | Kanungu | No | No* | No | Yes | 2 | No | Yes | Yes | Yes | 2 | Yes |

\* The machine was faulty and was last used 10 years back before TB-Speed intervention; CXR: chest X-ray

Regarding d-CXR set-up, in Uganda, in both DH, the X-ray department was renovated with the installation of lead doors and analogic X-ray machine was provided. DR plates was set-up on existing analogue radiography machines in 11 DH. One focal person from each country received an online training in setting up and using DR-plates by the manufacturer’s team in order to train radiographers at DH. All DH had at least 2 radiographers trained except in Rakai (Uganda) that had only one radiographer. Tablets, internet connectivity and electrical stabilisers were provided to all facilities (Table 2).

### Per-intervention implementation of d-CXR

#### Process and performance indicators of d-CXR

Of 3104 children with presumptive TB, 1642 (52.9%) had at least one d-CXR performed, of which 1505, 136 and 1 children had one, two and three d-CXR performed respectively, giving a total of 1780 d-CXR. Of them, 1773 (99.6%) were of good quality, and 1772/1773 (99.9%) were interpreted (Table 3).

**Table 3:**
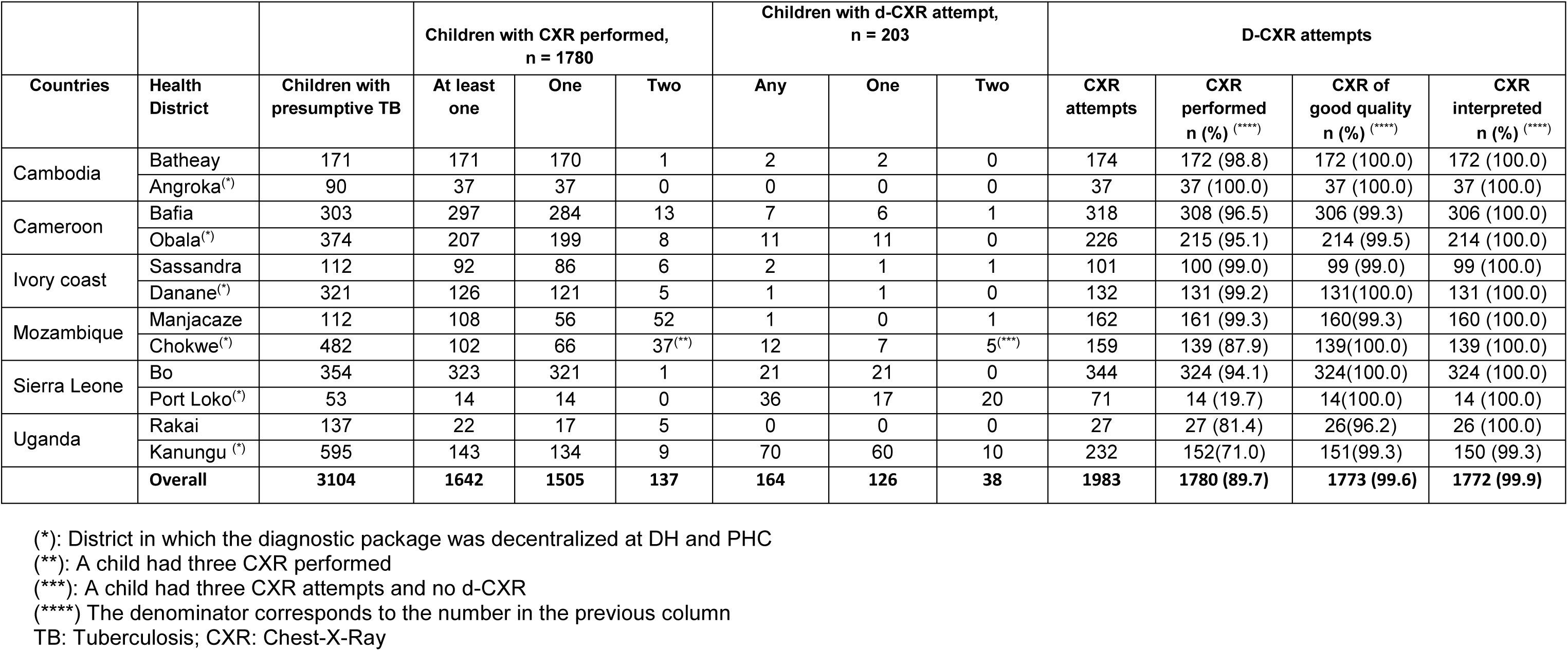
Cascade of care of chest X-ray in district hospitals per country, health district and overall.

A total of 46 sites supervision visits were performed in DH. The median score of the performance indicator assessing if d-CXR were performed as indicated was 5 in all countries, except Mozambique where it was 1. The median score regarding the process of loading of the d-CXR images was 1 in Mozambique and in Uganda, and the median score for the d-CXR interpretation indicator was 4 to 5 in all countries, except in Mozambique where it was 1 (Table 4).

**Table 4:**
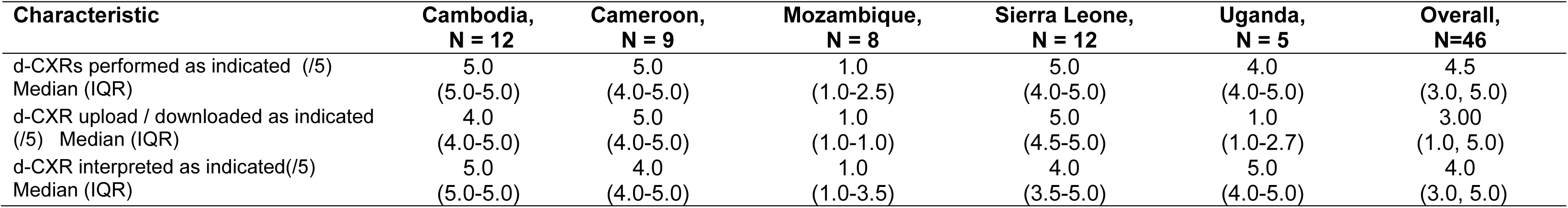
Median score results of the performance indicators from the site supervision visits in the district hospitals by country.

| Characteristic | Cambodia,<br>N = 12 | Cameroon,<br>N = 9 | Mozambique,<br>N = 8 | Sierra Leone,<br>N = 12 | Uganda,<br>N = 5 | Overall,<br>N=46 |
| --- | --- | --- | --- | --- | --- | --- |
| d-CXR performed as indicated (/5)<br>Median (IQR) | 5.0<br>(5.0-5.0) | 5.0<br>(4.0-5.0) | 1.0<br>(1.0-2.5) | 5.0<br>(4.0-5.0) | 4.0<br>(4.0-5.0) | 4.5<br>(3.0, 5.0) |
| d-CXR upload / downloaded as indicated<br>(/5) Median (IQR) | 4.0<br>(4.0-5.0) | 5.0<br>(4.0-5.0) | 1.0<br>(1.0-1.0) | 5.0<br>(4.5-5.0) | 1.0<br>(1.0-2.7) | 3.00<br>(1.0, 5.0) |
| d-CXR interpreted as indicated(/5)<br>Median (IQR) | 5.0<br>(5.0-5.0) | 4.0<br>(4.0-5.0) | 1.0<br>(1.0-3.5) | 4.0<br>(3.5-5.0) | 5.0<br>(4.0-5.0) | 4.0<br>(3.0, 5.0) |

#### HCWs’ perception, operational challenges and mitigation actions

HCWs interviewed valued the digital equipment provided, its technical features, the quality of images taken, and thereby being able to provide quality care to patients.

> *“What is working well is that, there are many ways it can be answered. The x-ray machine itself, because it gives services to different people, other patients and specifically the TB Speed children. Also, the imaging system what we call the DR system is also working well (…) It’s a sophisticated good machine.” (Uganda, Radiographer)*.

HCWs were happy with the quality of d-CXR they received. Very few reported issues on quality requiring to repeat the d-CXR.

> *“Technique of radiography is better though we follow up, we read the film x-ray through TB speed, most films have acceptable good quality.” (Cambodia, Nurse)*.

Some HCWs regretted the absence of printed X-rays when using d-CXR. They suggested that printed X-rays would be more easily accessible for HCWs for interpretation and would create more opportunities for learning among HCWs.

> *“Now, the chest x-ray they don’t produce film, yes after the x-ray, the doctor will go there and interpret the x-ray so I don’t have too have idea over it because I have not been there with the doctor to do the interpretation of the x-ray” (Sierra Leone, Nurse)*.

Regarding challenges, among reasons for not performing d-CXR reported in the clinical database, 44/203 (21.7%) were due to connectivity problem between the DR plate detector and the computer and 30 (14.8%) to electrical problems (Table 5). Difficulties in loading the d-CXR images, technical problems with the DR-plate and issues with internet connexion were also reported as common challenges in the supervision reports. During HCWs’ interviews, ensuring stable electricity was overall reported as one of the main challenges for d-CXR, occasionally resulting in system errors while performing X-ray due to the interruption of the internet connection. This often requires additional cost for the parents who need to come back the next day.

**Table 5:** Reasons for digital chest X-ray not performed by country.

|  | Cambodia<br>n=2 | Cameroon<br>n=19 | Ivory coast<br>n=4 | Mozambique<br>n=20 | Sierra Leone<br>n=78 | Uganda<br>n=80 | Overall n (%)<br>N=203 |
| --- | --- | --- | --- | --- | --- | --- | --- |
| Technical problem of the X-ray machine | - | 17 | 2 | 20 | 34 | - | 73 (35.9) |
| Problem of connectivity between the DR plates detector and the computer | 2 | 2 | - | - |  | 40 | 44 (21.7) |
| Non-specified technical issues | - | - | - | - | 34 | 6 | 40 (19.7) |
| Electrical power issues | - | - | 1 | - | - | 29 | 30 (14.8) |
| Absence of radiographer | - | - | - | - | 7 | 5 | 12 (5.9) |
| Administrative Issues | - | - | 1 | - | 3 | - | 4 (1.9) |

DR : Digital Radiography

> *I: What do you do then for example, child is coming and the electricity is off the whole day?*

> *R: I usually ask them whether they have, relatives close by where they can sleep so that the next day I take the x-ray. In any case, if they go back for them to come back it costs a lot to the client and TB Speed, if I ask and they have somewhere to sleep, then that is to their advantage of the parent. If they don’t have, I may admit them temporarily but if they cannot afford then they go back and choose another day to come back. (Uganda, Radiographer)*.

To mitigate the problems of internet, HCWs from DH often came to the radiography department to directly read the d-CXRs on the monitor. Refresher training of loading d-CXR, use of social media or platforms were recommended by supervisors to mitigate the problems of loading of the d-CXR images (Table S2). During HCWs’ interview, using social media tools for exchange d-CXR images or wireless telecommunication system were proposed as a mitigation action to the challenges with the loading of the image.

> *“Regarding result, we have system like Telegram, for me, I don’t have it but other staffs they have, my boss, and CXR film interpreter; for us, we don’t know how to interpret CXR. (Cambodia, Midwife)*

Many of the challenges raised by HCWs were not directly related to digitalization but to the X-ray machine breakdown, human resources’ absence and turn over, access issues (Table 5).

> *“The challenge is mainly around the distance and transport. But in most cases, the clients say they don’t have money to reach the hospital. …..” (Uganda, Nurse)*.

> *We have lost many patients because the patient will say I don’t have money to do the X-ray. (Sierra Leone, medical doctor)*.

## DISCUSSION

This study reports experience of implementing d-CXR for diagnosis of TB in children in high TB incidence and resource-limited countries at district level. Overall, the use of d-CXR resulted in a very high proportion of good quality CXR and this quality was very much appreciated by HCWs. These results show also the efforts required in terms of equipment, training and supervision to setup d-CXR at DH level and highlight the challenges faced during implementation and mitigation actions proposed by the end-users and the supervisors.

In term of site readiness, in our study settings, only 1 out of 12 district hospitals in six countries already had digital X-ray, reflecting the low coverage of digital X-rays in resource-limited countries at DH levels. Although not representative of DHs in all resource limited countries, pre-intervention findings highlight the level of investment and support that would be required for countries to setup digital X-ray at this level of health facility. However, this situation is expected to improve in the coming years with the introduction of mobile radiography and digital X-rays in the new Global Fund requests to increase access to radiography and ease the use of CAD currently recommended for TB screening in adults and under evaluation for diagnosis in children (17).

Regarding the challenges of using the d-CXR, the problems of connectivity between the DR plates captors and the computer was a common reason for the failure of d-CXR. The DR plates used in the study were chosen because of their rapidity between exposure and image acquisition but had the disadvantage of having fragile DR plate detector that could not support shocks and high temperatures, which may explain the dysfunctionalities. Manufacturers should consider designing less fragile sensors for markets of resource limited countries.

Even under study conditions, the process of loading images was challenging. This process can be complex and require good training and supervision as highlighted in the site supervision reports and good quality of the internet connection. Despite the providing of internet connection system to all DH during the d-CXR setup, several sites suffered from recurrent internet connection problems. To mitigate these issues, HCWs from DH had to come to the X-ray department to read the CXR on the monitor and others were using social medial tools for d-CXR exchange. This last solution is raising potential ethical issues related to the safeguarding of files and to the confidentiality in regards to the regulations in each country. Thus, when implementing digital X-ray, access rights, and handling of personal data should be organized using state of the art procedures such as encryption security, pseudonymisation and logging (18–20)

Another important challenge affecting the use of DR-plates was the frequent power shortages. A recent study of 72 health facilities from all health districts in Sierra Leone found that only 13 (18%) of rural facilities were connected to the national power grid. The authors pointed out that digitization of healthcare facilities should consider the use of renewable energy sources (21). Digitalization of X-ray at low level of healthcare would certainly require promoting the use of sustainable back-up systems for electricity, such as new wind or solar power.

The use of digitized X-ray requires a change in clinical practices, which may be challenging, as highlighted during interviews. There is still limited use of and experience in electronic data systems for healthcare at DH and many clinicians prefer having a printed CXR to be filed with the patient’s medical chart and carried during clinical rounds or shared with colleagues to discuss cases (22). It may indeed be challenging to incorporate a digitized tool requiring access to computers or tablets when patients’ files are still paper based. Digitized X-ray can be printed but this adds an additional cost and potential logistical and affordability issues.

Even if digital related issues can be solved, structural problems that affect access to X-ray were commonly reported to explain the lack of d-CXR and raised by HCWs during interview. Breaking down of the radiography machine was the first cause of d-CXR not performed in the study. The lack of suitable maintenance system is a common problem in many resource-limited countries that rely often on private contractors at a high cost(23). This raises questions about the sustainability of such interventions and the need to empower the health facilities to address most common technical issues with a back-up system for more complex ones.

In conclusion, d-CXR implemented at district hospital level in resource limited countries can provide good quality of CXR and could contribute to better diagnosis of childhood tuberculosis. It require significant investment in terms of equipment, training, maintenance and supervision that may be difficult to maintain under routine conditions, and consecutively a strong stakeholder engagement and buy in to ensure its sustainability (24). Hopefully with the upcoming CAD systems and t development of portable X-ray machines and their inclusion in the budget forecast to international funders, we could expect an increased access to digital X-ray in high TB burden and resource limited countries in the coming years (25).

## Data Availability

All data produced in the present study are available upon reasonable request to the authors

## ACKNOWLEDGEMENTS

We thank members of the TB-Speed Scientific Advisory Board who gave technical advice on the design of the study and approved the protocol. We thank all the healthcare workers of the participating health facilities as well as all TB-Speed staff in the field who coordinated qualitative study and colleagues from the TB Speed Decentralization study. We also thanks the Ministries of Health and national TB programs (NTPs) of participating countries; and the NTP district representatives who supported the TB-Speed Decentralization Study implementation.

The funding of TB Speed Decentralization study was provided by Unitaid.

## Authors’ contribution

J Orne Gliemann (JOG) conceptualized the qualitative study, JOG, B Joshi (BJ) and O Marcy (OM) supervised the qualitative study in the different countries at the central level. BJ and DM Massom (DMM) performed formal qualitative analyses, E Leroy- Terquem (ELT), PY Norval (PYN), conceived the training course with technical support from OM. JV Taguebue (JVT), A Salomao (AS). D Bunnet (DB), T Chhen Eap (TCE), L Borand (LB), C Khosa CK), R Moh (RM), J Mwanga-Amumpere (JMA), M Tan Eang (MTE), I Manhiça (IM), E Wobudeya (EW) and A Mustapha (AM) coordinated the implementation of the study in the different countries. B Fortune Melingui (BFM) collected the data and performed the statistical quantitative analysis with support from S Beneteau (SB) and M Bonnet (MB). BFM wrote the first draft of the manuscript with support from JOG and MB. All authors reviewed and approved the final version of the manuscript.

---

**Table S1:** Sociodemographic information of HCWs interviewed, TB-Speed Decentralization study, N=113, 2021.

| Characteristic | Cambodia,<br>n = 17 | Cameroon,<br>n= 24 | Ivory coast,<br>n= 16 | Mozambique,<br>n = 19 | Sierra Leone,<br>n = 20 | Uganda,<br>n = 17 | Overall,<br>N = 113 |
| --- | --- | --- | --- | --- | --- | --- | --- |
| Gender n(%) |  |  |  |  |  |  |  |
| Female | 8 (47) | 14 (58) | 2 (12) | 14 (74) | 16 (80) | 10 (59) | 64 (57) |
| Male | 9 (53) | 10 (42) | 14 (88) | 5 (26) | 4 (20) | 7 (41) | 49 (43) |
| Position n (%) |  |  |  |  |  |  |  |
| Medical doctor | 3 (18) | 5 (21) | 2 (12) | 3 (16) | 10 (50) | 2 (12) | 25 (22) |
| Midwife | 6 (35) | 0 (0) | 0 (0) | 0 (0) | 1 (5.0) | 0 (0) | 7 (6.2) |
| Nurse | 8 (47) | 19 (79) | 14 (88) | 16 (84) | 9 (45) | 13 (76) | 79 (70) |
| Radiographer | 0 (0) | 0 (0) | 0 (0) | 0 (0) | 0 (0) | 2 (12) | 2 (1.8) |
| Median Age (IQR) | 33 (30, 48) | 32 (30, 41) | 36 (33, 38) | 33 (28, 38) | 36 (31, 42) | 36 (33, 41) | 35 (31, 41) |
| Type of decentralization n (%) |  |  |  |  |  |  |  |
| DH Focused | 8 (47) | 13 (54) | 9 (56) | 10 (53) | 10 (50) | 7 (41) | 57 (50) |
| PHC Focused | 9 (53) | 11 (46) | 7 (44) | 9 (47) | 10 (50) | 10 (59) | 56 (50) |
| Type of health facility n (%) |  |  |  |  |  |  |  |
| District hospital | 6 (35) | 6 (25) | 3 (19) | 5 (26) | 6 (30) | 8 (47) | 34 (30) |
| Primary Health Centre | 11 (65) | 18 (75) | 13 (81) | 14 (74) | 14 (70) | 9 (53) | 79 (70) |

**Table S2:**
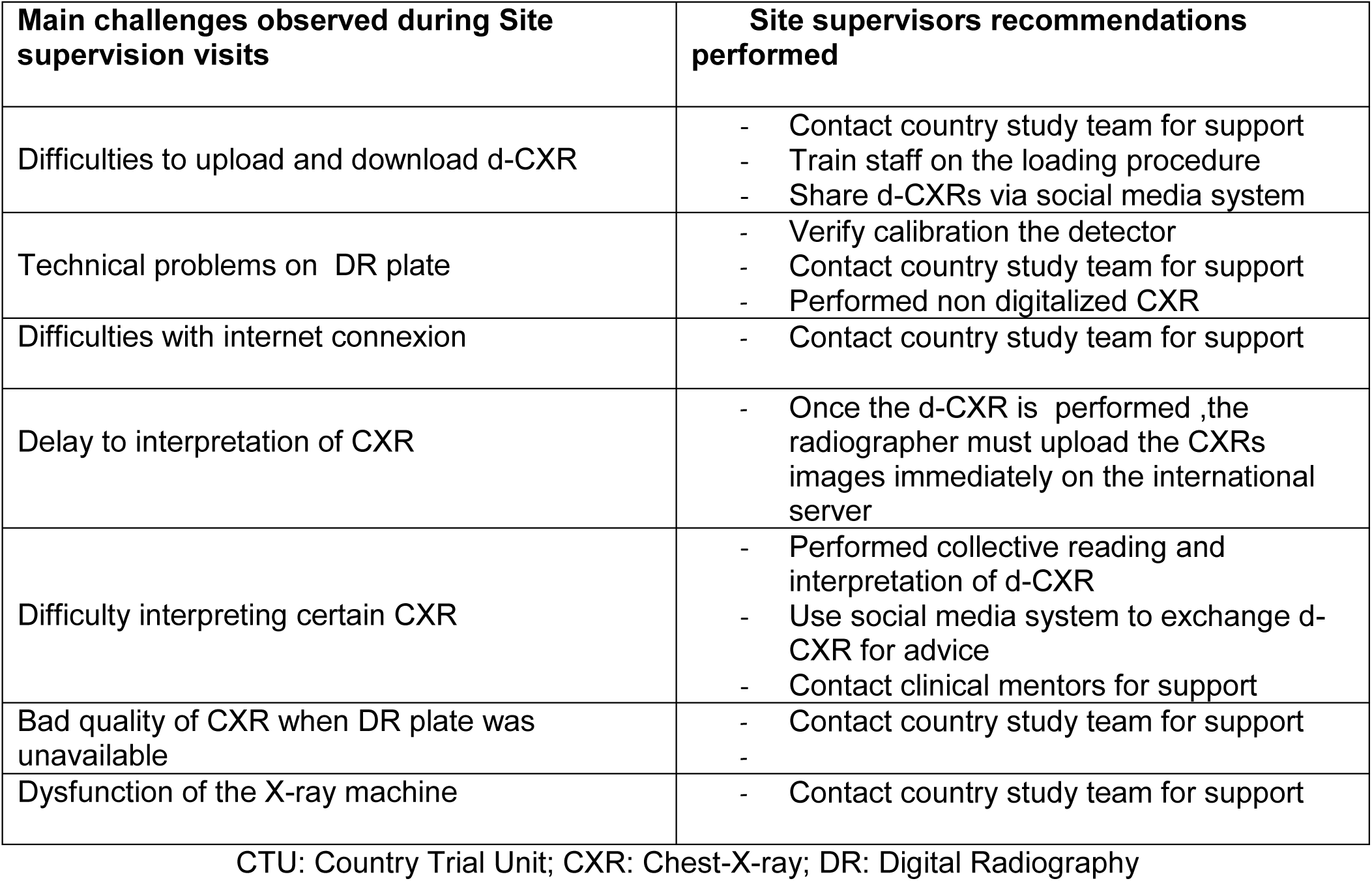
Challenges and recommendations identified from site supervision visits’ reports.

| Main challenges observed during Site supervision visits | Site supervisors recommendations performed |
| --- | --- |
| Difficulties to upload and download d-CXR | - Contact country study team for support<br>- Train staff on the loading procedure<br>- Share d-CXRs via social media system |
| Technical problems on DR plate | - Verify calibration the detector<br>- Contact country study team for support<br>- Performed non digitalized CXR |
| Difficulties with internet connexion | - Contact country study team for support |
| Delay to interpretation of CXR | - Once the d-CXR is performed ,the radiographer must upload the CXRs images immediately on the international server |
| Difficulty interpreting certain CXR | - Performed collective reading and interpretation of d-CXR<br>- Use social media system to exchange d-CXR for advice<br>- Contact clinical mentors for support |
| Bad quality of CXR when DR plate was unavailable | - Contact country study team for support<br>- |
| Dysfunction of the X-ray machine | - Contact country study team for support |

CTU: Country Trial Unit; CXR: Chest-X-ray; DR: Digital Radiography

